# Perinatal depression in Rural Kenya and the associated risk and protective factors: A prospective cohort study before and during the COVID-19 pandemic

**DOI:** 10.1101/2025.01.15.25320322

**Authors:** Caroline W. Wainaina, Joyce L Browne, Emmy Igonya, Fred Wekesah, Abdhalah K. Ziraba, Stephen Maina, Samuel Iddi, Estelle M. Sidze, Wendy Janssens, John De Wit, Kitty W.M. Bloemenkamp, Manasi Kumar

**Author notes:** Correspondent author: Caroline Wainaina.

## Abstract

**Purpose:** This study aimed to estimate the prevalence of perinatal depression in rural Kakamega, Kenya while exploring risk and protective factors in the context of the COVID-19 pandemic.

**Methods:** The mixed method approach employed i) quantitative data collected in a longitudinal maternal health evaluation conducted from October 2019 to May 2021 and ii) an ethnographic study conducted from March to July 2022, which provided detailed insights on the risk and protective factors of perinatal depression. The quantitative sample of 135 Pregnant and postpartum women was screened monthly for depression (>13) using the Edinburgh Postnatal Depression Scale (EPDS). Logistic regression assessed the association between socioeconomic status, clinical and psychosocial variables, and perinatal depression. A sample of 20 women was enrolled in the qualitative component of the study.

**Results:** The cumulative prevalence of perinatal depression was 11%. Depression symptoms were seen in 7% of pregnant women and 13% of mothers. During COVID-19, the odds of depression increased with maternal complications (aOR=7.05, 95%CI 1.66-29.94) and financial stress (aOR=1.40, 95%CI 0.66-2.98). Live birth outcomes reduced the odds of depression (aOR 0.03, 95%CI 0.002**-**0.73). Risk factors included health and healthcare challenges, lack of spousal and social support, intimate partner violence, and financial difficulties. Protective factors included adequate spousal and social support and access to economic resources, including digital platforms for soft loans and income hiding.

**Conclusion:** One in seven women experienced perinatal depressive symptoms. Increase in depression during the COVID-19 pandemic is indicative of the need for i) financial and social safety nets to cushion perinatal women during emergencies, ii) Integration of depression screening into healthcare and establishing confidential pathways for psychosocial support.

**What is already known about this topic - summarize the state of scientific knowledge on this subject before your study and explain why this study was necessary.:** Previous studies indicate that the prevalence of perinatal depression is rising in Kenya, with rates of antepartum depression ranging from 33% to 38% and postpartum depression between 19% and 27%. These studies mainly focus on urban and low-income populations. However, there is limited research on the burden of maternal depression in rural areas of Kenya.

**What this study adds - summarize the new insights gained from this study that were not previously known.:** This mixed-methods study provides valuable insights into the status of maternal depression in rural Kenya, marking the first household-based screening for depression conducted in such a setting. The findings reveal differences in the prevalence of depression between the periods before and during COVID-19. Additionally, the study details the risk and protective factors related to perinatal depression.

**How this study might affect research, practice, or policy - summarize the study’s implications.:** Understanding perinatal depression is essential for enhancing the integration of maternal mental health in both primary healthcare and community levels. Analyzing the risk and protective factors before and during the COVID-19 pandemic will provide insight into its impact on perinatal depression. The findings related to these factors will inform the development of targeted maternal health interventions.

## Introduction

Globally, around 322 million (4.4%) of the population suffer from depressive disorders, and at least 9% of those with depressive disorders live in Africa(World Health Organization, 2017). Perinatal depression is a common mental disorder that has its onset during pregnancy or up to one year postpartum(Gelaye et al., 2016). Perinatal depression (PD) is associated with adverse maternal outcomes, including adverse perinatal outcomes, maternal suicide, and suboptimal maternal-child bonding(Larsen et al., 2022). Perinatal Depression can also lead to adverse consequences for infants, including their physical health, psychological health, and development(Dennis & McQueen, 2009; Hurley et al., 2012; J et al., 2011).

A systematic review on perinatal depression in low- and middle-income countries (LMICs) found a pooled prevalence of 26.3% for antenatal depressive symptoms and 17% for postpartum depression (Dadi, Miller, et al., 2020; Dadi, Akalu, et al., 2020). In sub-Saharan Africa, antepartum depression is estimated at 14-24% in health facilities and 9.9% in rural Ghana (Adeoye et al., 2022; Getinet et al., 2018; Mwita et al., 2021; Weobong et al., 2014). Postpartum depression rates range from 7-27% in health facility screenings (Atuhaire et al., 2020). During the COVID-19 pandemic, the prevalence was estimated at 29% for pregnant women and 26% for mothers in the postnatal period (Caffieri et al., 2024).

Perinatal depression (PD) remains undetected and untreated in many low- and middle-income countries (LMICs) despite its increasing prevalence. Factors contributing to this evidence gap include stigma, inadequate education for healthcare workers and communities, insufficient investment in maternal mental health services, and poor implementation of screening and treatment guidelines. The lack of healthcare support and the impact of COVID-19 restrictions have further intensified the mental health burden on women.

Social determinants of perinatal depression include young maternal age, perinatal infections, complications, food insecurity, poverty, unemployment, lack of support, marital status changes, and intimate partner violence (Gelaye et al., 2016; Kumar et al., 2018; Osok et al., 2018). The COVID-19 pandemic worsened these risk factors by introducing fear of exposure, service disruptions, decreased healthcare utilization, stigma, reduced social interactions, and delayed detection of complications (Abrahams et al., 2022; Kotlar et al., 2021). These delays have led to higher rates of maternal deaths, morbidities, stillbirths, and perinatal depression, signaling further adverse effects on maternal and child health (Chmielewska et al., 2021; Slomian et al., 2019a).

In Kenya, most studies on perinatal depression focus on urban low-income populations, with estimated prevalence rates of antepartum depression ranging from 33-38%(Osok et al., 2018., Madeghe et al., 2021; Mochache et al., 2018) and 19-27% for postpartum depression(Kariuki et al., 2022; Ongeri et al., 2018). This emphasizes the need for empirical data in rural settings. Understanding the status of maternal depression is crucial for improving maternal mental health at primary healthcare and community levels. Analyzing risk and protective factors before and during the COVID-19 pandemic will highlight its effects on maternal depression. This study aims to estimate the prevalence of maternal depression in Kakamega, a rural county in Kenya, and identify associated factors in the context of the pandemic. We hypothesize a lower prevalence of perinatal depression in rural settings, as shown in the literature, with more factors categorized as protective rather than risk factors.

## Methods

### Study design

The quantitative component of the study was nested in a larger longitudinal evaluation study of the Innovative Partnership for Universal Sustainable Healthcare (i-PUSH) program in Kakamega, conducted between December 2019 and May 2021. Additional information on the i-PUSH study has been published elsewhere(Abajobir et al., 2020).

The ethnographic study conducted between March and July 2022 aimed to understand the factors associated with good or poor maternal mental health through the lived experiences of perinatal women from the i-PUSH study.

### Study setting

The study was conducted in the Khwisero sub-county of Kakamega County, a rural population in Western Kenya. Small-scale farming is the primary livelihood for the Khwisero community. The i-PUSH implementing partners, the PharmAccess Foundation and Amref Health Africa, selected the Khwisero area as the site for implementing the i-PUSH program.

### Study participants

#### Quantitative sampling

Women aged 15-49 who were pregnant or had a child under one year at the beginning of the i-PUSH study were recruited for this prospective study. A total of 135 women were enrolled in the cohort study and were followed up until their child was one year old, until they were lost to follow-up or the project ended. Women with new pregnancies during the follow-up period were also invited to participate, and they were screened for depression. Women who experienced an adverse pregnancy outcome (stillbirth, miscarriage, or spontaneous abortion) were not followed up for screening beyond the month when the outcome was recorded. This variation in enrolment was captured through repeated observations in our analysis and discussion of the findings.

#### Qualitative sampling

Women who were either pregnant or had a child under one year old were recruited into the participant observation study. The women were recruited from the i-PUSH sampling frame and were eligible if they had no to mild depression symptoms (scores of 0-12) during their last EPDS assessment in May 2021. We selected eligible participants (20 women) for the participant observation. The women were visited 8 times during the 4 months. The figure 1 below gives an overview of the quantitative sampling.

**Figure 1:**
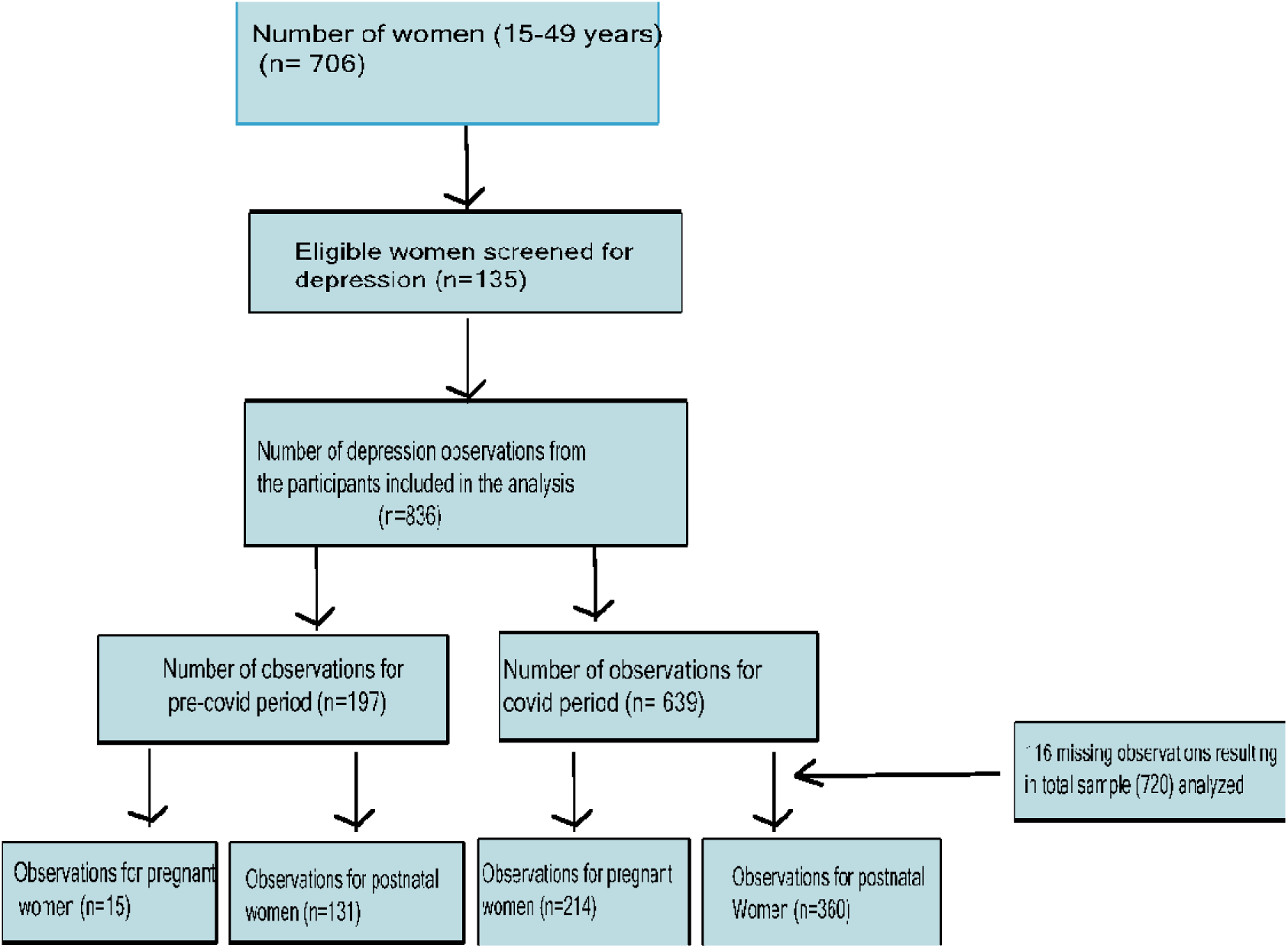
Overview of participants included for analysis.

### Assessments

Depression measure: We assessed depressive symptoms using the Edinburgh Postnatal Depression Tool (EPDS) from December 2019 to May 2021 for the quantitative sample and March to June 2022 for the qualitative sample. The EPDS consists of ten questions scored on a 5-point Likert scale, categorized as no to mild depression (0-12), moderate (13-19), and severe (20-30). For analysis, we categorized the outcome as binary: no depression (cut-off <13) and depression (cut-off >13). The questionnaire was available in Kiswahili and English based on participants’ preferences. The translation and validation of the Kiswahili version of the EPDS have been published elsewhere(Linnet Ongeri, 2015). The assessment was done monthly during the project period.

Socio-demographic information: We collected participant characteristics, including age, marital status, employment, perceived health status, parity, and education, at baseline measurements and transformed these variables into categories for analysis.

Clinical factors: We collected data on pregnancy, maternal and neonatal complications, pregnancy outcome (live birth or not), and place of delivery (home or health facility)using a survey questionnaire.

Psychosocial factors: The controlling behavior variable comprised five questions measuring freedom of movement, socializing, and women’s decision-making. It was used as a proxy measure of intimate partner violence. The data was collected during baseline measurements. The financial stress of households was also evaluated by asking about worries about not having enough food due to a lack of money or fear of loss of livelihood, including the effect of the COVID-19 pandemic. The data was collected monthly during the COVID period.

#### Data analysis Quantitative analysis

The data was analyzed using Stata version 15. Misra et al.’s (2003) perinatal mental framework guided the analysis. The framework describes the factors influencing an individual’s health and well-being. We adapted the framework to include the postpartum phase, which exhibits similar pathways to subsequent short—and long-term outcomes that can affect maternal and infant health and functioning.

The proximal factors directly impact a person’s health, such as controlling behavior, financial stress, and clinical characteristics. The socio-economic characteristics represent the distal factors/determinants that affect the individual.

In the study, maternal and neonatal complications are classified as risk factors for perinatal depression. They are viewed as proximal factors, with their short-term effects likely heightening the risk of perinatal depression.

Descriptive statistics were calculated for sociodemographic characteristics (maternal age, marital status, education, and employment status), clinical factors (pregnancy outcomes, pregnancy, and postnatal complications, and skilled birth deliveries), and psychosocial factors (controlling behavior and financial stress) against the outcome of interest (depression).

The multivariable regression model identified variables uniquely contributing as risk or protective factors for the outcome (postnatal depression). We focused on postpartum depression only for the bivariate and multivariable analysis because there was no pre-COVID outcome data (depression scores) for pregnant women and low numbers of the outcome during the COVID period (n=17/229). The analysis was done by grouping the data into the pre-COVID period (December 2019-Mid March 2020) and the Covid period (March 2020-May 2021).

*Qualitative component:* Two field research assistants documented their observations while” deep hanging out” with the participants during their daily activities. The ‘deep hanging out’ provides deep engagement and a glimpse into the participants’ lives (Greetz, C., 1998). Prompts were used to engage the participants in informal conversations about their daily lives and their views on different issues affecting them. The conversations were conducted in Kiswahili or the common language in the area (Luhya). The audio recordings and field notes were transcribed. The observation checklist, generated through prior coding by a team of five researchers, informed the development of the codebook framework. The field notes were then coded using NVivo 10 software through a content thematic analysis approach. (Braun, V & Clarke, 2006) The consolidated checklist for reporting (COREQ) was applied in this paper(Tong A, Sainsbury P, 2007).

### Ethical consideration

Ethical approval was secured from Amref Health Africa’s Ethical and Scientific Research Committee (AMREF ESRC P1119/2021), along with permissions from the County Government of Kakamega and a research permit from NACOSTI. Trained research assistants conducted the consent process, informing participants of their rights, the study’s objectives, and the use of their information. Participants who agreed signed the informed consent form.

## Results

During the study, 135 eligible women were screened at least once for depression. We analyzed their depression data using repeated measures. Of this sample, 46 were assessed during the pre-COVID period, and 89 were assessed during the COVID period. Figure 1 shows the sample selection for analysis.

The mean age of the women was 26 years (SD 5.89). 67% were married, while 33% were unmarried, separated, or divorced. Most had primary education (68%), with 32% having secondary education or higher. Half had more than two children, and 34% had one child. Less than a third (29%) were employed, and 95% belonged to the Luhya ethnic tribe.

The cumulative prevalence of perinatal depression (PD) was 11% (n=82/720). Depressive symptoms were found in 7% (n=17/229) of pregnant women and 13% (n=65/491) of mothers. Depression during pregnancy was only reported at 8% (n=17/214) during the COVID-19 period. In the postnatal period, pre-COVID-19 rates were 11% (n=15/131), and during COVID-19 were 14% (n=50/360). Due to the limited observations during pregnancy, we focused on the postnatal data.

Table 1 below shows the association between the proximal and distal factors and the presence of depressive symptoms in the participating women during pre-COVID and COVID-19 periods. Under the participant characteristics, the study found that depressive symptoms were more common among women aged 24-34, 18% (13/71) and 35 years and over, 21% (14/52) during the pre-COVID and COVID periods, respectively. Lower education (primary) and higher parity(>2) were associated with depression at 16% (17/107) and 14% (17/104), respectively, during the pre-COVID period. Having adverse pregnancy outcomes such as stillbirth, miscarriage, spontaneous abortion, and maternal and neonatal complications were significantly associated with depressive symptoms. Women in controlling relationships had a positive association with depression during the COVID period at 16% (39/243).

**Table 1:** Bivariate analysis of associated factors vs postpartum depression.

|  | Precovid |  | P-value | Covid |  | P value |  |
| --- | --- | --- | --- | --- | --- | --- | --- |
|  | No depression | Depression |  | No Depression | Depression |  | Total (n) |
| **Age** |  |  |  |  |  |  |  |
| *15-24 year* | 96(97) | 3(3) | 0.00* | 269(92) | 392(47) | 0.01* | 392(47) |
| *24-34 year* | 58(82) | 13(18) |  | 247(88) | 351(42) |  | 351(42) |
| *35+* | 24(89) | 3(11) |  | 52(79) | 93(11) |  | 93(11) |
| **Marital status** |  |  |  |  |  |  |  |
| *Never Married* | 72(92) | 6(8) | 0.45 | 175(91) | 270 (33) | 0.23 | 270(33) |
| *Married* | 106(89) | 13(11) |  | 385(88) | 557(67) |  | 557(67) |
| **Education** |  |  |  |  |  |  |  |
| *Primary* | 107(86) | 17(14) | 0.02* | 380(88) | 554(68) | 0.57 | 554(68) |
| *Secondary/Tertiary* | 66(97) | 2(3) |  | 170(90) | 257(32) |  | 257(32) |
| **Employment status** |  |  |  |  |  |  |  |
| *No* | 128(92) | 11(8) | 0.18 | 375(90) | 556(68) | 0.15 | 556(68) |
| *Yes* | 48(86) | 8(14) |  | 172(86) | 256(32) |  | 256(32) |
| **Parity** |  |  |  |  |  |  |  |
| *No children* | 25(100) | 0(0) | 0.00* | 53(91) | 83(12) | 0.47 | 83(12) |
| *1 child* | 55(98) | 1(2) |  | 158(88) | 235(34) |  | 235(34) |
| *More than 2 children* | 87(84) | 17(16) |  | 225(86) | 366(54) |  | 366(54) |
| **Perceived health status** |  |  |  |  |  |  |  |
| *Poor/Average* | 49(83) | 10(7) | 0.02* | 175(89) | 256(31) | 0.976 | 256(31) |

Note:

|  |  |  |  |  |  |  |  |
| --- | --- | --- | --- | --- | --- | --- | --- |
| *Good/Very good* | 129(93) | 9(7) |  | 385(89) | 571(69) |  | 571(69) |
| **Pregnancy outcome** |  |  |  |  |  |  |  |
| *No live births* | 0(0) | 2(100) | 0.00* | 3(100) | 5(1) | 0.49 | 5(1) |
| *live births* | 116(90) | 13(10) |  | 307(88) | 486(99) |  | 486(99) |
| **Maternal complications** |  |  |  |  |  |  |  |
| *No* | 89(91) | 8(9) | 0.16 | 233(90) | 356(73) | 0.00* | 356(73) |
| *Yes* | 27(82) | 6(18) |  | 77(76) | 134(27) |  | 134(27) |
| **Neonatal complications** |  |  |  |  |  |  |  |
| *No* | 111(92) | 10(8) | 0.01* | 294(86) | 463(95) | 0.94 | 463(95) |
| *Yes* | 5(63) | 3(38) |  | 13(87) | 23(5) |  | 23(5) |
| **Pregnancy complications** |  |  |  |  |  |  |  |
| *No* | 104(88) | 14(12) | 0.65 | 292(86) | 456(93) | 0.55 | 456(93) |
| *Yes* | 12(92) | 1(8) |  | 18(82) | 35(7) |  | 35(7) |
| **Place of delivery** |  |  |  |  |  |  |  |
| *No health facility* | 46(88) | 6(12) | 0.98 | 70(85) | 134(27) | 0.84 | 134(27) |
| *Health facility* | 70(86) | 9(11) |  | 238(86) | 355(73) |  | 355(73) |
| **Controlling behavior** |  |  |  |  |  |  |  |
| *Low to moderate controlling* | 81(91) | 8(9) | 0.68 | 207(90) | 319(49) | 0.05* | 319(49) |
| *Most controlling* | 74(89) | 9(11) |  | 204(84) | 326(51) |  | 326(51) |
| **Financial Stress** |  |  |  |  |  |  |  |
| *No financial stress* |  |  |  | 250(91) | 275(33) | 0.27 | 275(33) |
| *Financial stress* |  |  |  | 496(88) | 561(67) |  | 561(67) |

Significance level established at ( $p < 0.05$ ) with a Confidence interval of 95%

The adjusted odds ratio (aOR) of having maternal complications was significantly associated with postpartum depression aOR 33.5 (95%CI 0.95-1182.45) and aOR 7.1 (95%CI 1.66-29.94) during pre-COVID and COVID periods respectively. Being employed was significantly associated with postnatal depression aOR 5.4 (95%CI 1.12-26.3) during the COVID period. Being in a controlling relationship was strongly associated with postnatal depression aOR 3.2 (95%CI 0.16-64.84) and AOR 2.6 (95%CI 0.59-11.01), during pre-COVID and COVID respectively, though the results were not statistically significant in our study. The pregnancy outcome variable (live birth vs. no live birth) was omitted from the model because of collinearity with neonatal complications (miscarriage, abortion, and stillbirth).

**Table 2:**
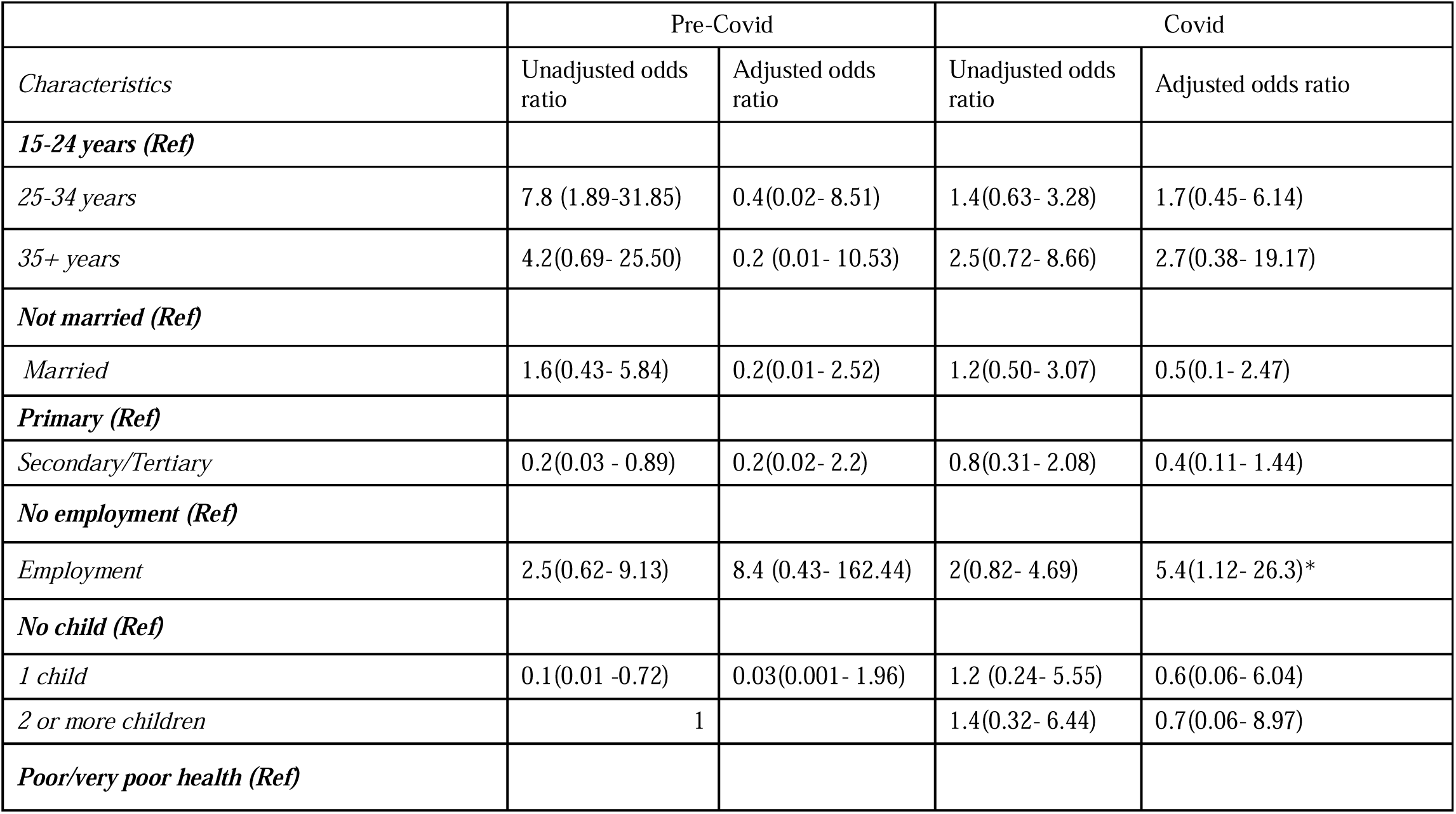
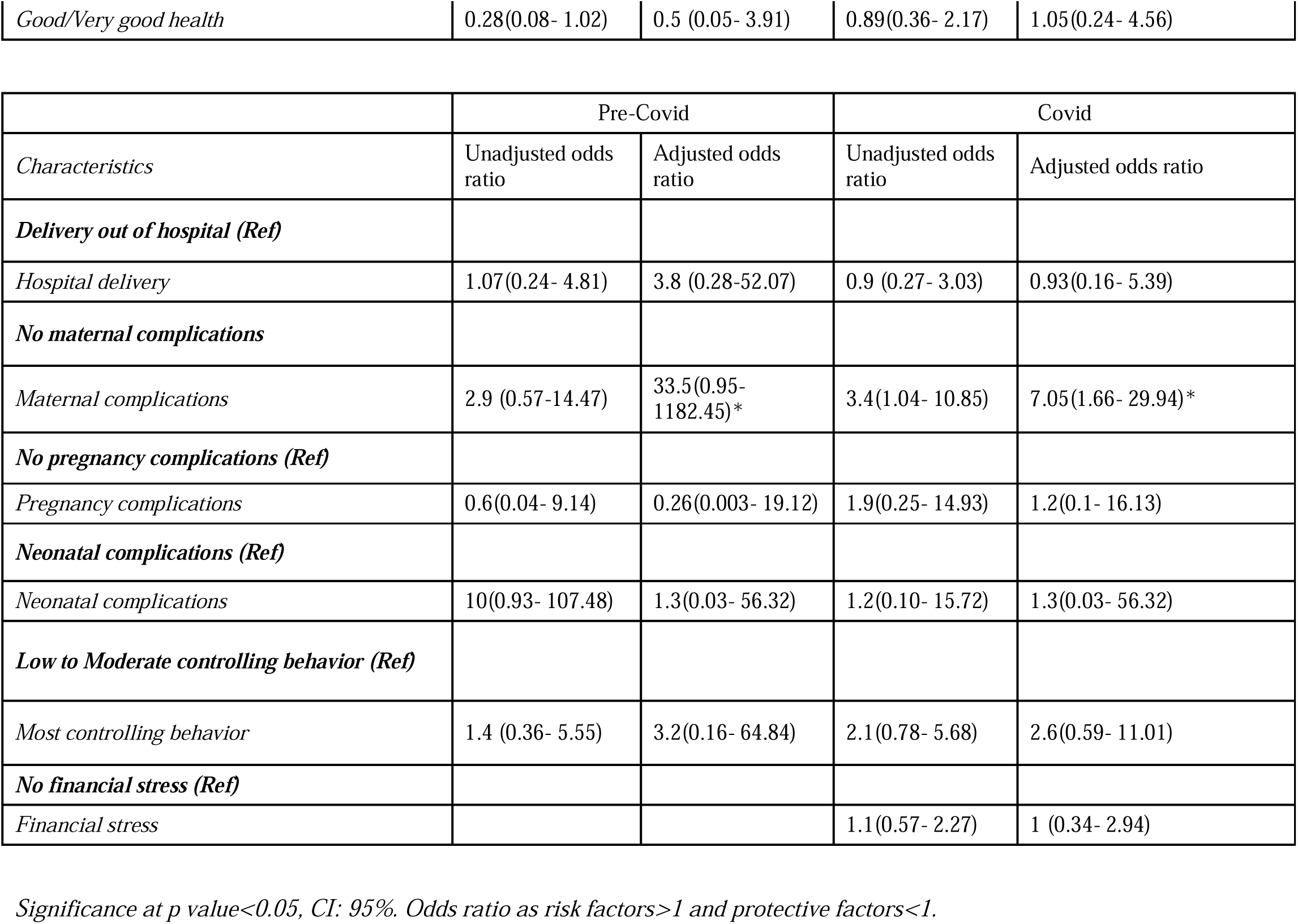
Multivariate analysis of covariates against postnatal depression.

### Qualitative findings

#### Participant characteristics

Most women were aged between 25 and 30 years (n=12/20), 9 were pregnant, and 11 had a child younger than one year. Most had primary-level education, and half were unemployed.

**Table 3:**
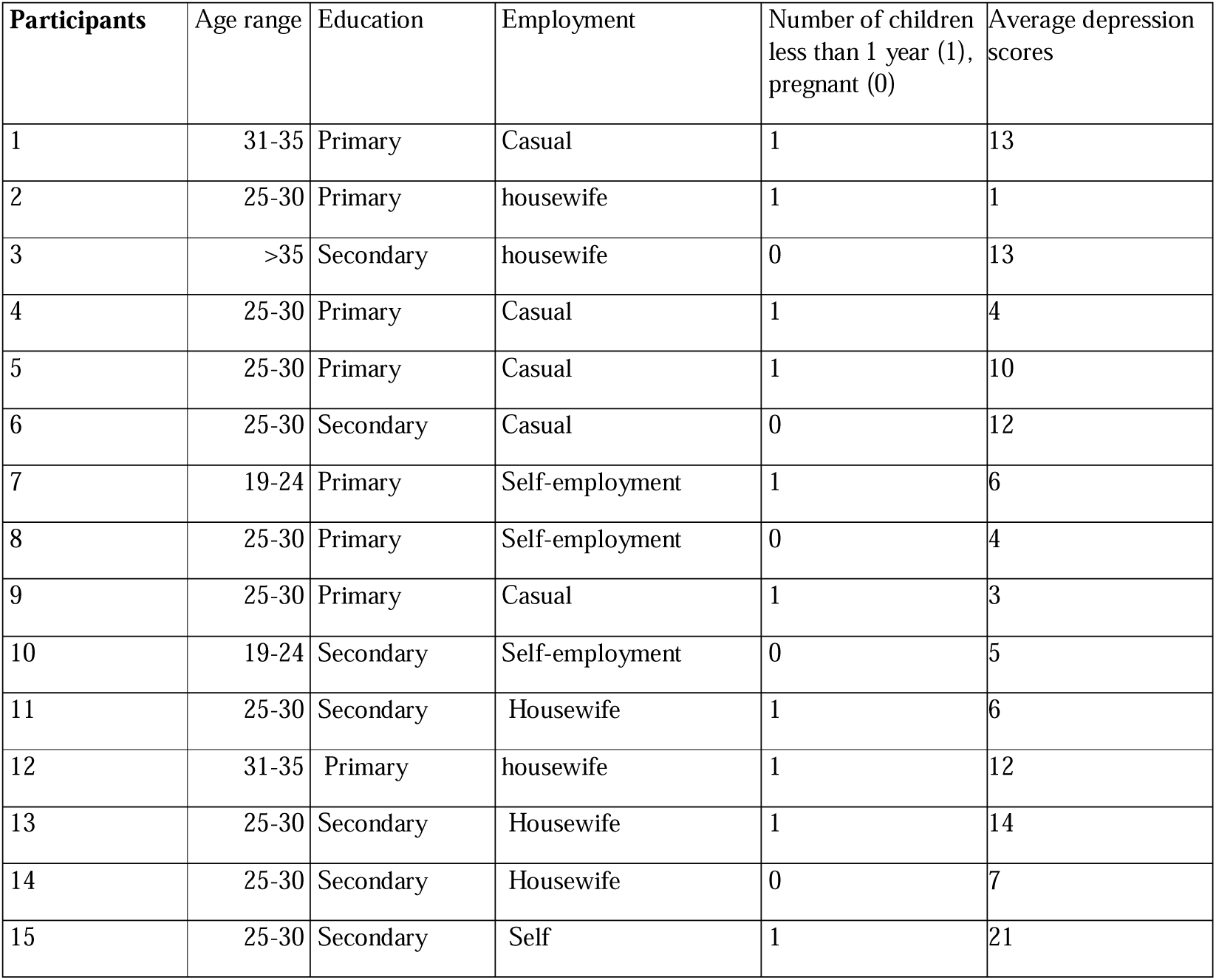
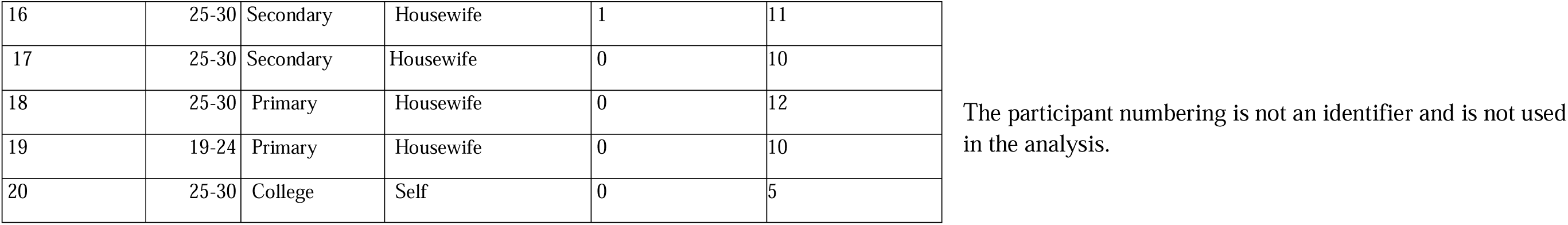
Characteristics of participants for the participant observation exercise.

We deductively identified four themes and categorized them as risk factors based on narration from the qualitative participants. The themes are highlighted in Table 4 below. The participant numbering is not an identifier.

**Table 4:** Identified themes and subthemes.

| Associated factors | Theme | Sub-theme |
| --- | --- | --- |
| Risk factors | Pregnancy and motherhood journey | Unplanned/unintended pregnancies |
|  |  | Health and healthcare challenges |
|  |  | Food insecurity |
|  | Poor Social support/relationships | Inadequate spousal support |
|  |  | Poor relationship with in-laws |
|  | Poor financial access | Challenges in accessing economic resources |
|  | Intimate partner violence/controlling spouses | Verbal, physical abuse, controlling behaviour |
| Protective factors | Spousal support | Emotional, physical, and financial support |
|  | Functional social support | Emotional, physical, and financial support |
|  | Access to economic resources | Access to platforms for saving and borrowing |
|  |  | Income hiding |

### Risk factors identified

#### Difficult p3regnancy and childbirth phase

##### Unplanned/unintended pregnancies

A few women described having unplanned or unintended pregnancies that had led to elevated levels of stress. One of the reasons for the unplanned pregnancies was failed family planning methods; another reason was financially constrained relationships.

> *“By the way, I had not expected to get pregnant again. I was using family planning, and I still conceived… I no longer trust the family plan methods (Participant 3)*.

##### Health and healthcare challenges

Women indicated going through health challenges during the perinatal period. The illness, fatigue, and restlessness affected their self-confidence and limited their productivity in the home and community.

> “*[..] I have been feeling severe fatigue most of the time. That is why I am unable to wake up very early in the morning” (Participant 10)*.

Access to maternal healthcare faced challenges such as high costs for ultrasound scans and medications. Some women struggled to deliver at health facilities due to complications or long travel distances. One participant shared a negative experience, blaming healthcare workers for her prolonged labor, and noted that the absence of a doctor for an emergency cesarean led to complications, resulting in her baby’s death six days after delivery.

> *” I am very sure if the health care workers acted promptly, they could have saved my child…during my ANC visits, I found their services were just good, but on that night of delivery, I regret why I went there”. (respondent 10)*

##### Food insecurity

Care and support during the perinatal period are critical to women’s health and well-being. The provision of food is essential during this period. Food insecurity led to women feeling stressed, compounded by their inability to provide for themselves.

> *“I am always in deep thought. I wish I were not pregnant because I could withstand these conditions (food scarcity) well. However, now that I am pregnant and need to eat frequently, yet there is not enough food, it worries me a little bit” (Participant 2)*.

Food insecurity also stressed women who were breastfeeding. Women linked their inability to breastfeed adequately to a lack of appropriate and adequate food.

#### Social support/relationships

##### Spousal support

Advanced pregnancy and childbirth forced women to depend on their spouses for financial support. However, the spouses, who often work as motorcycle riders, farmers, or small business owners, often struggled to provide a stable income. This led to an inability to adequately meet the household’s basic needs, such as food, school fees, and healthcare.

> *“The impact we have felt in terms of food. He (the woman’s spouse) has reduced the amount he spends on food. (Participant 2, pregnant)*

Women also encountered stress when they were unable to fulfill their financial obligations, as one narrates below:

> *"I am really stressed because I need money to send to my parents… but I depend on my husband, and he has let me down”. (Participant 8 with a young child).”*

Alcohol abuse was prevalent in the study area attributed to harsh economic times, leaving men incapable of providing for their families, exacerbating the women’s mental stress.

##### Relationship with in-laws

A lack of social support from women’s in-laws contributed to stress during the perinatal period. The women mentioned mistreatment and abuse by in-laws, including interfering with their economic activities;

> “*He (in-law) could tell me in front of my customers that I should know that his cows are more important than my customers. So, you find that most of my customers started boycotting coming here for my services” (Participant 8)*.

Another participant narrated how her in-laws had mistreated her because of having male children. The issue she indicated was the availability of land to inherit. Inheritance in African cultures is given to males (sons), and as such, the more males there are, the more demand there is for land to inherit(Bau & Fernández, 2023).

> *“There was a time when my in-law used to harass me a lot. It reached a point when he told me to pack all my belongings and leave his homestead because I was giving birth to boys only…asking where I would take the male children now that there is not enough piece of land to accommodate them” (Participant 5)*.

#### Poor financial access

The post-COVID economy faced challenges like business closures and financial stress. Women highlighted how pregnancy and childbirth limited their economic opportunities. Financial independence allowed them to engage in community activities and women-saving groups like table banking. Without this participation, they experienced stress and missed social and economic support.

> “*That is another source of stress; I have missed two sittings without making any contributions (financial). If I could have money and attended chama, at least I could have requested them to give me a soft loan to take this child to school” (participant 8)*.

#### Intimate partner violence/controlling spouses

Some women faced intimate partner violence (IPV), including physical, verbal, and emotional abuse, often stemming from men’s alcoholism or trust issues, particularly with financially independent women. Some contemplated leaving or even suicide, while a few stayed due to fear or stigma.

> “*he insulted me…he pushed me to the extent of contemplating committing suicide”* (Participant 5). “*she says that (seeking counseling) will bring trouble to her, and the spouse will abuse her if he knows”…*(Participant 13).

#### Protective factors

##### Spousal support

Amidst a lack of income, women indicated receiving financial support, though irregular, from their spouses, which alleviated their immediate financial stress.

> “*I have just been surviving on the financial support received from my husband. He sent me some money through Mpesa (mobile money) twice (Participant 8).”*

One participant shared how her spouse supported her during the loss of their newborn. The spouse provided both physical and emotional support throughout her grieving process.

##### Functional social support

Some of the women interviewed also cited family support as a coping strategy. They relied on their paternal and maternal families for help when they could not fend for themselves.

> “*… I do ask for help from my relatives, such as my parents and relatives. They have been very supportive. My relative sent me money to buy food…(Participant 9)*

In addition to family, other communities and the church have supported the participants in overcoming their challenges and building resilience.

##### Access to economic resources

Access to economic resources allowed women to meet basic needs and save for emergencies. They used digital platforms, such as mobile apps, for soft loans and to manage their savings. Table banking saving groups (Chama) allowed women to save money and access loans. By saving, they became eligible for loans or received contributions when it was their turn.

> *I received the merry-go-round (savings group) contributions…I gave it to my spouse to buy the baby’s needs and food. Saving is good. The money came in at the right time when I was about to deliver.” (Participant 2)*

##### Income hiding

Some women save or receive money secretly because their spouses would withhold funds or misuse their money, especially those without phones who rely on their spouses for digital access.

> *“If I have to tell my husband…I will say a lower figure than what I received…my spouse does not know I save” (Participant 13)*.

## Discussion

In the study conducted in rural Kenya among perinatal women who were between 15-49 years, the prevalence of perinatal depression was 11%. Antepartum depression (APD) was only reported during the COVID period at 8%. Postpartum depression (PND) was 11% vs 14% during the pre-COVID and COVID period, respectively. Discussions on mental stress triggers helped women identify challenges during pregnancy and motherhood, such as complications, lack of support, limited finances, and intimate partner violence. Protective factors included positive support and access to financial resources. Differences in prevalence were observed between pre-COVID and COVID periods, likely due to increased risks.

Government measures like lockdowns and curfews affected rural areas in Kenya. Women expressed that the closure of churches and schools deprived them of emotional and spiritual support. The population noted that the economic situation worsened during the pandemic. The perceptions and behaviors during the pandemic have also been supported by research from other rural populations. Research shows that during the pandemic, perceptions and behaviors varied significantly between rural and urban populations(Greteman et al., 2022). For instance, a study in urban Kakamega (Paul et al., 2022) found that women doubted the reality of the pandemic and did not follow restrictions, highlighting minimal differences between pre-COVID and COVID times. Over time, populations tend to normalize and adjust to the situation, resulting in decreased mental health incidents (Husain et al., 2023).

In contrast, another study (Paul et al., 2022) conducted in urban Kakamega, the area close to the study site, indicated that women did not believe the pandemic was real and, therefore, did not adhere to the restrictions. This perception from an urban population near the rural study area illustrates the minimal difference between the pre-COVID and COVID periods. The temporal changes in the effect of the pandemic on mental health demonstrate that over time, populations normalize and adjust to the new reality, leading to decreased mental health incidents (Husain et al., 2023).

The prevalence of postpartum depression in low- and middle-income countries ranges from 17% to 20% (Dadi, Miller et al., 2020; Gelaye et al., 2016). A study in Siaya and Homa Bay Counties reported 16.8% postpartum depression among HIV-negative women, compared to 24.5% for antepartum depression (Larsen et al., 2022). Overall, low- and middle-income countries have higher rates of perinatal depression than high-income countries, with a systematic review indicating a 12% prevalence and 17% incidence of postpartum depression (Howard & Khalifeh, 2020). In rural areas, prevalence rates range from 9.9% in Ghana (Weobong et al., 2014) to 23% in Ethiopia (Lodebo et al., 2020).

The lower antepartum prevalence in our rural population may be due to women’s normalization of risk factors, like being in controlling relationships, especially for those financially dependent on their spouses. Research shows a higher rate of postpartum depression compared to antepartum, consistent with this study’s findings. Kendell et al. (1987) noted that women are 22 times more likely to experience depression after childbirth than during pregnancy.

Our multivariate regression analysis found a strong link between maternal complications and depression (Dadi, Akalu et al., 2020; Slomian et al., 2019b). Women delivering at local health facilities showed a higher likelihood of developing depression. While other studies indicate that access to healthcare reduces maternal complications and improves survival rates (Joseph et al., 2016), factors like facility deliveries, mode of delivery, mistreatment, fear of childbirth, and delivery outcomes can increase mental stress for women.(Räisänen et al., 2013; Silveira et al 2019).

Our qualitative research provided in-depth insights that complemented the quantitative findings on risk and protective factors related to mental stress. Notably, women faced health and healthcare challenges that contributed to their stress levels. A systematic review highlighted a strong link between poor health and increased rates of depression among outpatients compared to the general population (Leight et al., 2022). Quantitatively, financial stress—specifically the inability to afford food—was identified as a risk factor for perinatal depression. Qualitative results revealed that financial dependence created stress, prompting women to seek economic opportunities to enhance their mental well-being. Previous research highlights poverty and low socio-economic status as significant risk factors for perinatal depression (Gelaye et al., 2016; Dadi, Miller et al., 2020; Madeghe et al., 2021). Although we did not directly inquire about intimate partner violence, we used controlling behavior as a proxy and found that depressed women were more likely to be in controlling relationships. One study noted the link between powerlessness and perinatal mental health in a rural Ethiopian population.

Spousal and family support are vital for women’s mental well-being during the perinatal period. Male involvement helps women cope with stress, making their participation important from pregnancy to motherhood. In cases of marital conflict, maternal family relationships can provide protective support against abuse. Studies link strong social networks to increased resilience against perinatal depression (Saligheh et al., 2014).

Due to restrictions during the COVID-19 pandemic, social isolation mainly affected urban areas. However, a study in India found that close-knit rural families provided essential social support, contributing to improved mental health during the pandemic (Husain et al., 2023). Thus, enhancing the resilience of perinatal women to mental health challenges requires focusing on strong social networks and women’s empowerment.

Due to challenging economic conditions, employment, including self-employment and casual labor, emerged as a risk factor during COVID-19. Economic empowerment is essential for reducing gender inequality, lowering postpartum depression, and improving social welfare participation (Leight et al., 2022; Madeghe et al., 2021).

### Strengths and Limitations of the Study

#### Strengths

The study used high-frequency household data to track individual depression trends in a rural setting. A cohort study design helped control for confounding participant characteristics. We employed the validated Edinburgh Postnatal Depression Scale Tool (EPDS), including a locally translated version for better comprehension. By combining quantitative and qualitative methods, we provided descriptive data alongside insights into participants’ experiences, highlighting risk and protective factors. The findings are vital for policy and program implementation, offering real-time mental health trend data for targeted interventions.

#### Limitations

This study’s limitations include a small sample size (n=135), which limited analyses and reduced statistical power. Monthly screenings may have led to rehearsed answers, and the concealment of pregnancies resulted in fewer pregnant participants. Additionally, women were enrolled at different times, preventing some from receiving the full 18 months of observation, which affected the total observations and depression prevalence rates. The short recall period may have also missed some depressive symptoms outside the screening timeframe.

## Conclusion

The prevalence of maternal depression in rural areas is a significant issue. Our findings on postpartum depression rates align with those in Western Kenya and the global average, highlighting the need for depression screenings in maternal health services. Antepartum depression rates were lower than average.

Factors such as maternal complications, controlling behavior, financial stress, and facility childbirth were exacerbated by the COVID-19 pandemic, indicating broader social influences on women’s mental health during the perinatal phase. To tackle this, maternal health interventions should improve healthcare systems, promote respectful care, and enhance social support. Regular screenings for perinatal depression in households and health facilities are crucial for early treatment and prevention of severe outcomes.

## Acknowledgments

The study was conducted in Khwisero, Kakamega County. We thank all participants, the field researchers, and the County Government for their support.

## Contributions of the authors

Conceptualization-Caroline Wainaina, Emmy Igonya, Joyce Browne, Manasi Kumar, Estelle Sidze, Wendy Janssens, Frederick Murunga Wekesah, John De Wit and Kitty Bloemenkamp; Formal analysis Wainaina, Stephen Maina, Abdhalah Ziraba, Samuel Iddi, Joyce Browne, Manasi Kumar; Supervision of data collection: Caroline Wainaina, Emmy Igonya, Estelle Sidze, Wendy Janssens; Writing of first draft-Caroline Wainaina; Writing and reviewing of the paper-All the authors.

## Patient and public involvement

The public was not involved in the design, conduct, reporting, or dissemination of the study.

## Data availability statement

The quantitative data can be obtained from the African Population and Health Research Center’s microdata portal at https://microdataportal.aphrc.org/index.php/admin/catalog/edit/167.

The qualitative data is currently not publicly available but might be available to the corresponding author upon reasonable request.

